# Potential associations of selected polymorphic genetic variants with COVID-19 disease susceptibility and severity

**DOI:** 10.1101/2024.03.13.24304197

**Authors:** Orsolya Mózner, Edit Szabó, Anna Kulin, György Várady, Judit Moldvay, Vivien Vass, Andrea Szentesi, Ágoston Jánosi, Péter Hegyi, Balázs Sarkadi

## Abstract

In this study, we analyzed the potential associations of selected laboratory and anamnestic parameters, as well as 12 genetic polymorphisms (SNPs), with clinical COVID-19 occurrence and severity in 869 hospitalized patients. The SNPs analyzed by qPCR were selected based on population-wide genetic (GWAS) data previously indicating association with the severity of COVID-19. We confirmed the associations of disease with several clinical laboratory and anamnestic parameters and found an unexpected association between less severe disease and the loss of smell and taste. In most cases, selected SNP analysis supported earlier results by indicating genetic associations with hospitalization and disease severity, while the potential role of some previously unrecognized polymorphisms has also been observed. A genetic association was indicated between the presence of a reduced-function ABCG2 transporter variant and a less severe disease, which was also observed in diabetic patients. Our current results, which should be reinforced by larger studies, indicate that together with laboratory and anamnestic parameters, genetic polymorphisms may have predictive value for the clinical occurrence and severity of COVID-19.

## Introduction

The course of the COVID-19 pandemic has shown great individual variability, from asymptomatic infection or mild disease to severe disease, in some cases leading to patient death. In addition to direct viral toxicity, hyperreactivity of the immune system may significantly worsen patients’ conditions, and endothelial damage, microvascular injury, and hypercoagulability with thrombosis may also occur. In addition to the signs of severe pulmonary disease, the poor prognosis of hospitalized patients is predominantly indicated by laboratory data reflecting an overreaction of the immune system to viral infection. The already established factors affecting the severity of acute COVID-19 include older age, overweight status, and comorbidities (e.g., diabetes) [1–4]. The currently used pharmacological agents to treat COVID-19 have questionable effects in severe cases, and positive results are mostly observed at the initial stage of this disease (see)[5–9]).

Increasing amounts of data are available about the potential effects of individual genetic backgrounds on the course and severity of COVID-19. Genetic factors affecting disease severity include molecular switches of the cellular immune response, e.g., the double-stranded RNA sensor Toll-like receptor 3 (TLR3) and type I interferon (IFN)-related pathways (see[10–14]). Rare mutations, such as loss-of-function deletions in the genes coding for the participants of these pathways, causing deficiencies in the expression of TLR3, TICAM1 (Toll Like Receptor Adaptor Molecule 1), IRF3 and IRF7 (Interferon Regulatory Factors 3 and 7), and IFNAR1 and IFNAR2 (Interferon Alpha and Beta Receptor Subunits 1 and 2), have been described as genetic factors underlying severe pneumonia in patients with COVID-19 [11, 12].

At the population level, rare mutations may not affect the clinical development of acute or chronic COVID-19. In contrast, the presence of certain single nucleotide polymorphisms (SNPs) and related complex haplotypes (a series of coinherited SNPs) is closely associated with the clinical course of COVID-19 in GWA studies (see [14]), especially regarding severe lung inflammation and the consequently required ICU treatment. Interestingly, several such haplotypes originate from the genetic material of the Neanderthal man, as segments (haplotypes) of the Neanderthal genome with characteristic SNPs are still present in the human genome. The Neanderthal haplotypes are almost absent in African populations (at the origin of modern humans) but present with variable frequencies in various other geographical regions (see [15, 16]).

One of these Neanderthal haplotypes is located on chromosome 3, with a large number of coinherited (haplotype) SNPs (chr3p21.31 – lead SNP: rs73064425), and the region includes six genes *(SLC6A20*, *LZTFL1*, *CCR9*, *FYCO1*, *CXCR6* and *XCR1),* which are potentially important in the immune response to this viral disease. The *CCR9, CCR6 and XCR1* genes encode cytokine receptors, which may be involved in the “cytokine storm” in COVID-19 patients. The LZTFL1 protein (encoding chr3, p21.31 – lead SNP: rs73064425) regulates ciliary transport processes in the airways and is potentially an important factor in the treatment of COVID-19 [17–24]. As reported in genome-wide association studies (GWAS), the presence of this Neanderthal-related haplotype correlates with doubling of the occurrence of severe respiratory disease in COVID-19 patients [15, 16].

Another Neanderthal-originating gene fragment, suspected to correlate with the severity of COVID-19, is a haplotype within the dipeptidyl peptidase 4 *(DPP4-DT)* gene on chromosome 2 (q24.2) with the leading SNP rs117888248/rs118098838. The DPP4 protein, together with ACE2, was found to be one of the binding sites of the SARS-CoV-2 virus, and the presence of the *DPP4-DT* Neanderthal-related haplotype in a heterozygous form was reported to double the appearance of severe COVID-19, while in a homozygous form, it correlated with a quadruple occurrence of severe disease [15, 16, 25, 26].

Other Neanderthal-related haplotypes reported to correlate with the severity of COVID-19 include genetic elements coding for the DPP9 protein (chr19, p13.3 – lead SNP: rs2109069) and the interferon alpha receptor (IFNAR2) protein (coding chr21, q22.1 – lead SNP: rs2236757) (see refs [27, 28]). In OAS1, the OAS2 and OAS3 protein coding regions of a haplotype (chr12, q24.13; lead SNP: rs10735079) were reported to correlate with less severe disease, while the protection against severe disease conferred by the Neanderthal *OAS* locus was substantially lower than the increased risk conferred by the chromosome 3 locus (refs [15, 16]). These interferon-induced OAS proteins produce short polyadenylates from RNA based on their ribonuclease activity and have antiviral effects in virus-infected cells.

In addition to the genes related to this Neanderthal heritage site, some relatively frequent SNPs in membrane receptor or transporter proteins may also have significant effects on acute or long-term COVID-19. The ACE2 membrane protein is the key receptor for SARS-CoV-2 binding to its cellular targets, and a haplotype in the *ACE2* gene (chrX, p22.2 – lead SNP rs2285666) has been shown to reduce the expression of this receptor. The variable presence of the T/A and G/C alleles may contribute to malaria sensitivity [29] and has been implicated in affecting COVID-19 susceptibility [30]. Similar to *ACE2*, *RAVER1* (chr191, p13.2– lead SNP: rs74956615) is a risk factor for COVID-19 infection [17, 18, 31]. Mucin encoded by *MUC5B* (chr11, p15.5 – lead SNP: rs35705950) is an important component of the innate immune response, and the presence of its promoter polymorphism, rs35705950, has a positive effect on the outcome of lung diseases through increased expression of MUC5B [32–34]. Loss of sense of smell (anosmia) or taste (ageusia) are characteristic symptoms of COVID-19 and are the earliest and most frequently reported indicators of the acute phase of SARS-CoV-2 infection. These symptoms have been reported to be variably associated with recovery, and the polymorphism of *UGT2A1* (chr4, q13.3– lead SNP: rs7688383) is one of the most significant genetic markers associated with loss of smell and taste [35, 36].

In addition to SNPs that have been previously studied in the context of COVID-19, our research also focused on genetic variations in membrane transporter proteins known to play key roles in cellular processes. Genetic polymorphisms in the PMCA4b protein, which widely affect cellular calcium homeostasis (a minor haplotype (referred to as “*ATP2B4*.haplo1”) in the regulatory region of the corresponding *ATP2B4* gene, chr1, q32.1 - lead SNP: rs1541252) (see [37]), or SNPs within GLUT1, a key cellular transporter protein of glucose and vitamin C (encoded by the *SLC2A1* gene, chr1, p34.2 - lead SNP: rs1385129 [38]), may modulate disease severity by affecting general metabolism. A frequent polymorphic genetic variant of the ABCG2 transporter (chr4, q22.1 – SNP rs2231142, resulting in a Q141K amino acid change) reduces uric acid, xenobiotic, and drug transport in various tissues and tissue barriers [39, 40].

While genome-wide association (GWA) studies are useful for exploring the role of potential genetic factors in large populations, small effects increasing disease risk in many cases cannot be distinguished from background noise, and only genetic variants with strong statistical evidence can be considered significant. Therefore, targeted molecular genetic studies should help to establish firm and relevant connections between selected polymorphic variations and the course of COVID-19, which affects numerous tissues and organs.

In this work, based on existing (although at that time mostly preliminary) data, we selected and analyzed 12 haplotypes and lead SNPs (see Supplementary Materials, Table 1) potentially relevant to the reaction of the human body to this viral disease in 869 hospitalized patients in Hungary. At the time of data collection (between 2020 and 2021), the dominant SARS-CoV-2 variants were the original Wuhan variant and the Delta variant, and there was no effective vaccination or treatment for this disease.

Although only a limited number of potentially relevant genetic variants were detected in a relatively small number of patients, the present study may help to decipher the role of several clinical, anamnestic and genetic parameters in the occurrence and severity of this viral disease. Additionally, when extended to a larger number of patients, our results may provide a personalized tool for assessing the expected course, severity, and long-term, chronic effects in COVID-19 patients.

## Results

### 1. Clinical parameters - potential association with disease severity

We collected and analyzed data from 869 hospitalized COVID-19 patients. For the statistical analysis, disease severity was grouped into two main categories: 592 patients were regarded as moderate (originally labeled 195 mild and 397 moderate), and 277 patients were regarded as severe (originally labeled 141 severe and 136 critical). Among the examined patients, 465 were male and 404 were female. The mean age at the time of study for severe and moderate patients was 65 and 61 years, respectively. A total of 274 hospitalized patients had diabetes mellitus; among these patients, 18 had type I diabetes, 252 had type II diabetes (92%), and 594 did not have diabetes. Fifty-three patients had bronchial asthma, while 815 patients did not. A total of 168 patients reported a loss of taste, and 602 patients had no such symptoms. The loss of a sense of smell was reported by 164 patients, while no such symptoms were recorded in 605 patients. The overall analysis of these data, as well as the genetic parameters related to their H-W distributions and the Khi^2^ probe, are presented in the Supplementary Materials, Table 3.

The large amount of anamnestic and clinical data for COVID-19 patients treated at reporting clinics requires a detailed and systemic statistical analysis that correctly reflects the potential effects of these variables on the severity of the disease. This analysis will be performed in an independent publication, while Figure 1A represents an initial statistical analysis.

**Figure 1.**
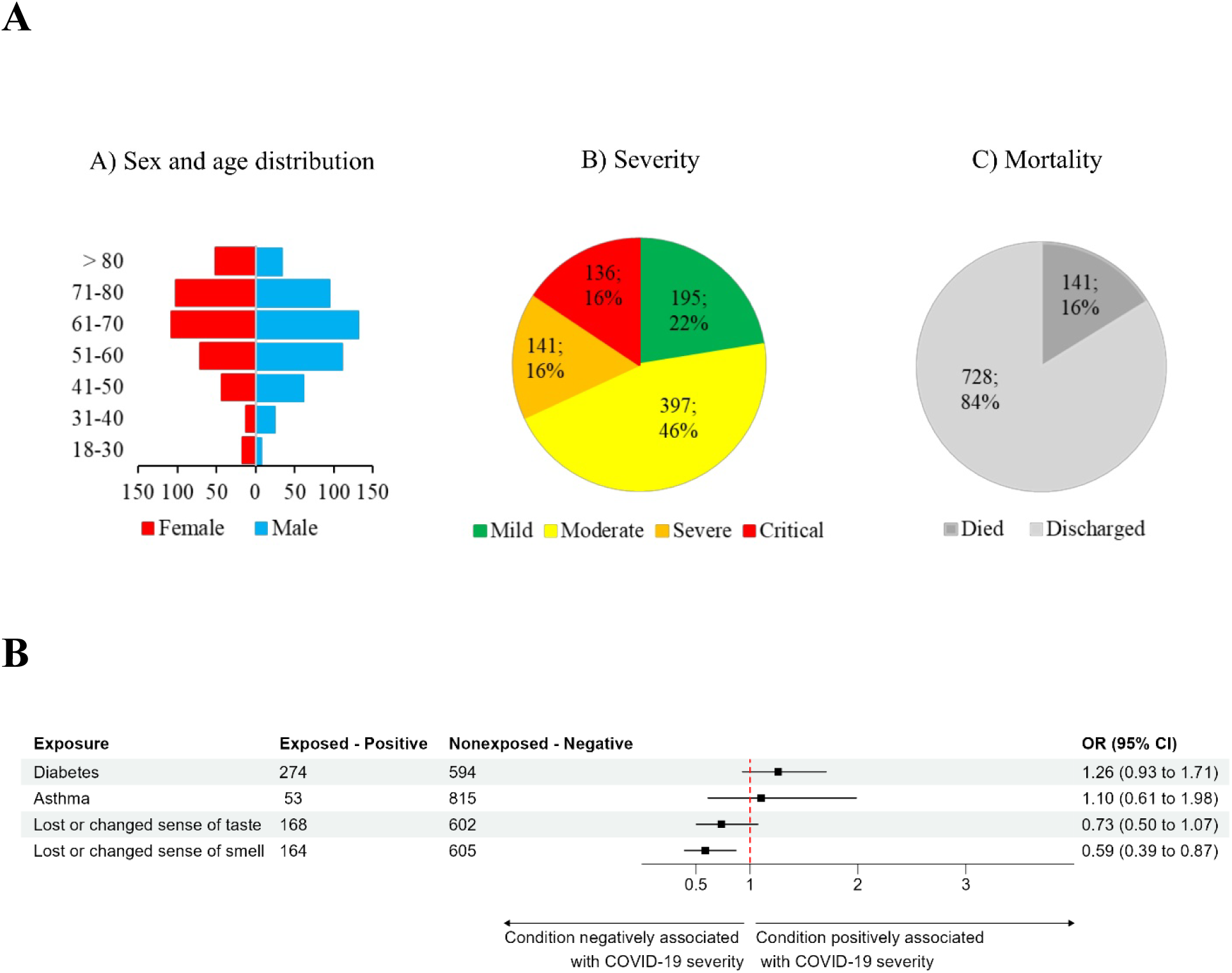
A. Summary of some key data for the population and disease severity of the COVID-19 patients included in this study. Figure 1B. Associations of selected anamnestic parameters (categorical values) with disease severity in COVID-19 patients. The figure provides the number of positive and negative patients for each parameter and shows a forest plot for the odds ratios (ORs) for the associations of the selected parameters with disease severity. The forest plot indicates either lower average or higher average OR values and the respective 95% confidence intervals for severe, as compared to the mild COVID-19 cases.

Figure 1B. shows the statistical analysis for some categorical clinical parameters with respect to disease severity. Here, we selected anamnestic parameters that have already been indicated in the relevant literature to affect COVID-19 severity and had statistically acceptable numbers of patients in this study.

As indicated in Figure 1B, in accordance with the literature, we found a major effect of preexisting diabetes (in 92% of patients type 2 diabetes) on the severity of COVID-19 at the clinic. In contrast, we did not find a significant effect on disease severity in patients with asthma, while the potential effects of high blood pressure or cancer were not analyzed in this work. This lack of statistical significance may be caused by the relatively low number of patients suffering from these conditions. Interestingly, we found a significant association between the loss or altered sense of smell (and a similar tendency in the case of the loss or change of taste) and a less severe clinical form of the disease.

### 2. Potential associations of selected genetic polymorphisms with hospitalized patients with COVID-19 and disease severity

In this analysis, we first examined the minor allele frequencies (MAFs) of the polymorphic variants of the selected genes in all the hospitalized COVID-19 patients and compared these MAF values to those in the representative European population. Our results are presented in conjunction with data from the 1000 Genomes Project and the ALFA database for comparison. Due to occasional discrepancies in European MAF values between these two major sources, we included both in our analysis for a comprehensive evaluation. As shown in Table 2, we found that in certain cases, these MAF values in the hospitalized patients showed major differences compared to the general European MAF values.

Although the two databases for the mean European MAF values are somewhat different (especially in the cases of *RAVER1* and *MUC5B*), these data together indicate that individuals carrying the minor variants of *LZTFL1* and *RAVER1* (and possibly *ATP2B4* and *MUC5B*) are generally more susceptible to hospitalization-requiring COVID-19 than the general population. In contrast, those carrying minor variants of the *IFNAR2* gene (and probably also *OAS3)* are significantly less likely to have a hospitalization-requiring disease.

In the following, for each selected polymorphism, we investigated whether the presence of the given polymorphism (minor variant) is associated with COVID-19 **severity** in hospitalized patients.

In our analysis, we examined the MAF values of polymorphic variants in selected genes **among hospitalized COVID-19 patients with severe disease** and compared these MAF values with those in the representative European population, as reported in the databases (see Supplementary Figure 1).

These data show that individuals carrying minor variants of *LZTFL1,* possibly *RAVER1* and *MUC5B,* are generally more likely to have severe COVID-19 than the general population. In contrast, those carrying minor variants of the *IFNAR2* gene (and probably also *OAS3)* are significantly less likely to have severe COVID-19.

Next, we analyzed the number of hospitalized COVID-19 patients carrying minor genetic variants (heterozygous+homozygous forms) and wild-type variants for each parameter and calculated the OR for **associations between the selected parameters and disease severity**. In this analysis, we compounded the patients carrying the heterozygous and homozygous minor variants (labeled “minor variant”) into one group. The overall results are shown in Figure 3 in the form of a forest plot for the calculated OR.

**Figure 2.**
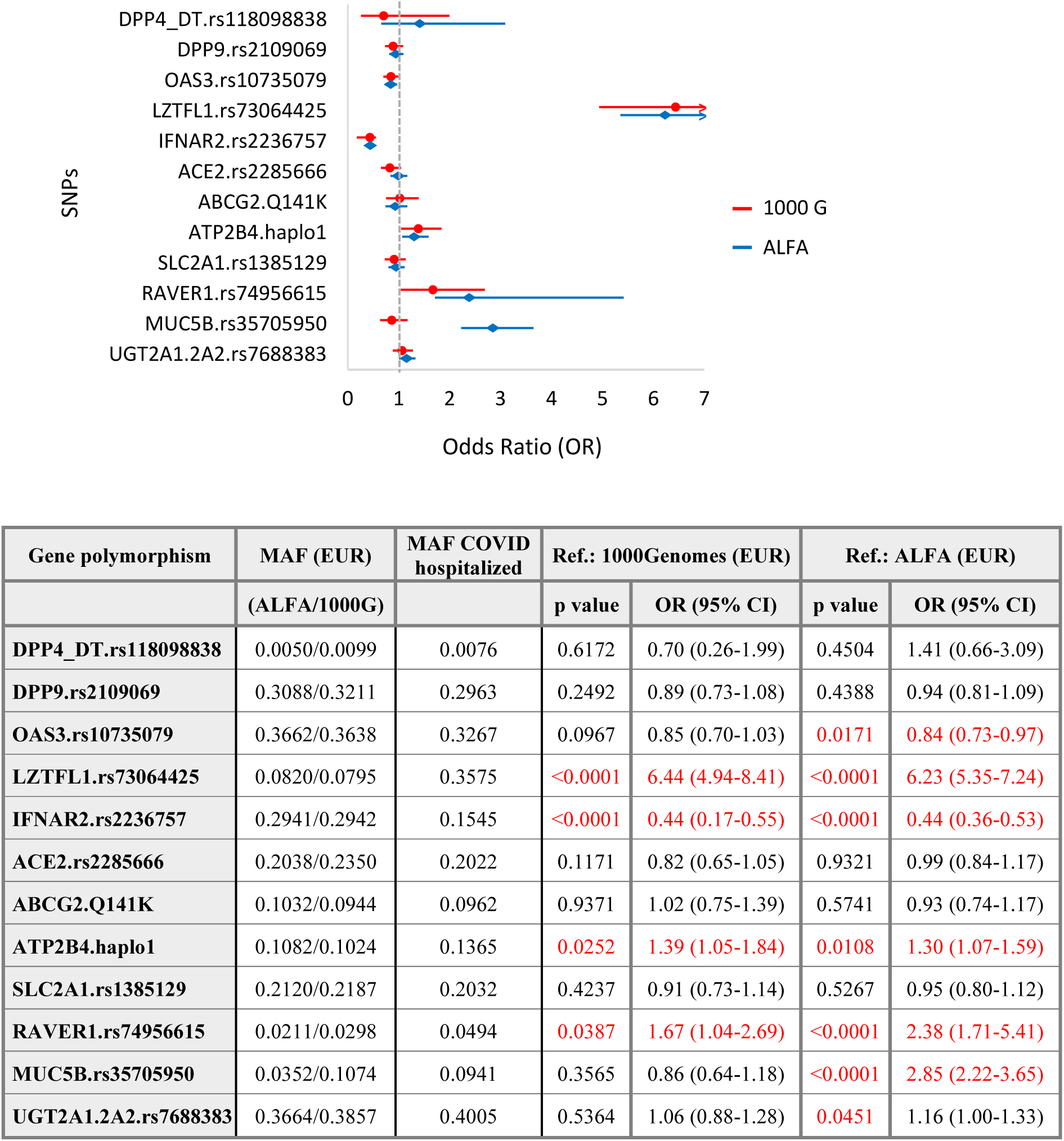
Allele frequencies of minor variants (MAFs) in the general population and hospitalized COVID-19 patients. Odds ratios and significance.

**Figure 3.**
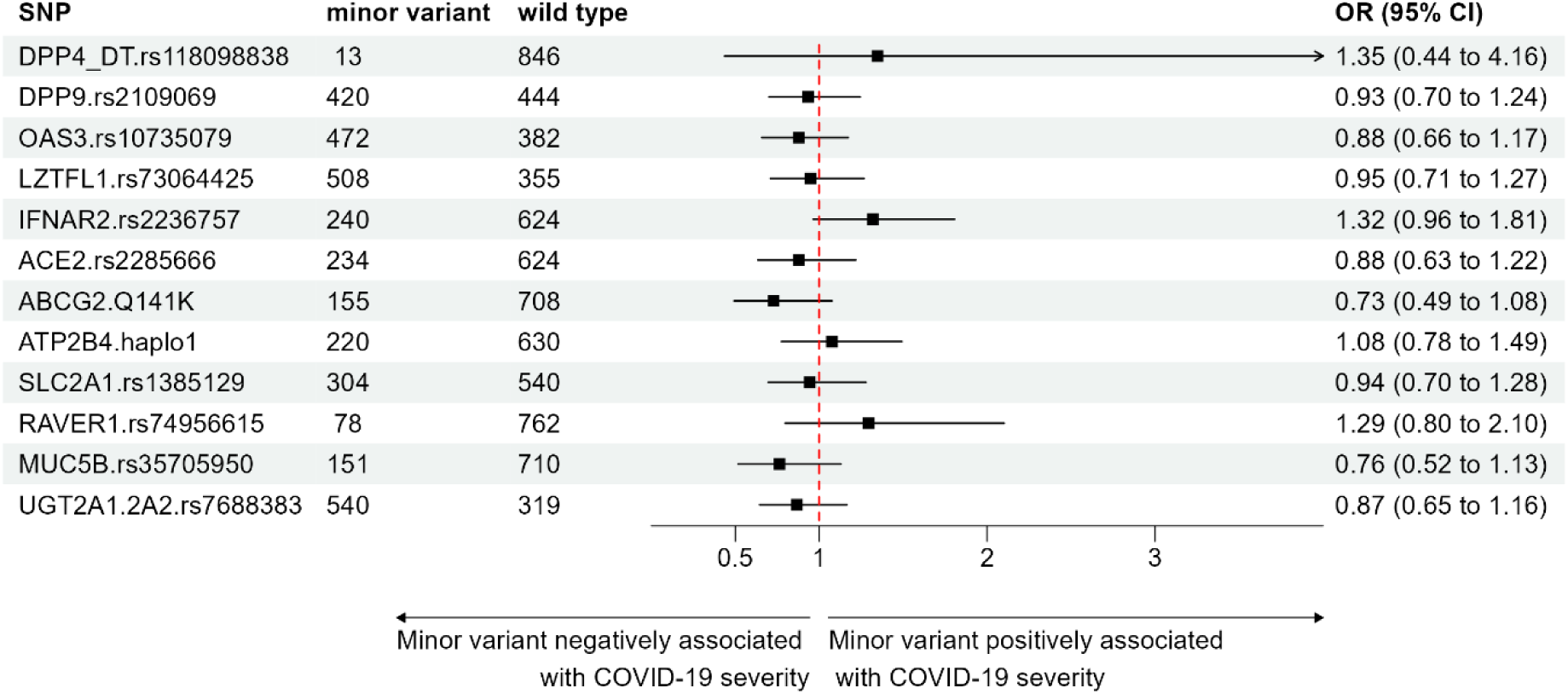
Associations between selected polymorphic genetic variations and disease severity in COVID-19 patients. The figure provides the number of patients carrying the minor genetic variant (heterozygous+homozygous forms) and the wild-type variants for each parameter and shows a forest plot for the odds ratios (ORs) for associations of the selected parameters with disease severity. The forest plot indicates either lower or higher average OR values than 1 and the respective 95% confidence intervals for patients with severe COVID-19severe, as compared to the mild COVID-19 cases.

As shown in Supplementary Figure 1, in the case of the presence of minor variants in the genes *IFNAR2* and *RAVER1*, we found an indication of the more severe form of COVID-19 in the hospitalized patients. In contrast, this analysis revealed a less severe form of the disease in the presence of minor variants of the *ABCG2* and *MUC5B genes*. As indicated by the bars in the figure, none of these differences reached statistical significance; that is, the 95% confidence intervals did not entirely exclude the null value (OR=1). Nevertheless, even with this relatively low number of patients, these data indicate a potential association between disease severity and these genetic polymorphisms.

### 3. Potential association of genetic polymorphisms with preexisting type 2 diabetes mellitus (T2DM) in COVID-19 patients

According to the literature and our data shown above (see Figure 1.), the presence of diabetes in patients correlates with greater COVID-19 severity. Therefore, we analyzed the potential associations of the examined genetic polymorphisms with the presence of diabetes and disease occurrence and severity.

First, we analyzed the MAF values of the polymorphic variants of the examined genes by comparing the diabetic hospitalized patients to the mean European minor allele frequencies (see Supplementary Figure 2).).

These data indicate that patients with type 2 diabetes carrying minor variants of *LZTFL1* and *RAVER1* (and probably also *MUC5B)* are more likely to have hospitalization-requiring COVID-19 than the general population. In contrast, diabetic patients carrying minor variants of the *IFNAR2* gene (and probably also *ABCG2)* are less likely to have severe COVID-19.

Next, we analyzed the associations of the minor variants of the polymorphisms in the 12 genes examined with COVID-19 **severity in diabetic and nondiabetic** patients (Figure 4). As shown in the figure, due to the relatively small number of patients, the confidence intervals for the potential associations of disease severity with preexisting diabetes and the examined genetic polymorphisms were large in most cases, and these differences did not reach statistical significance.

**Figure 4.**
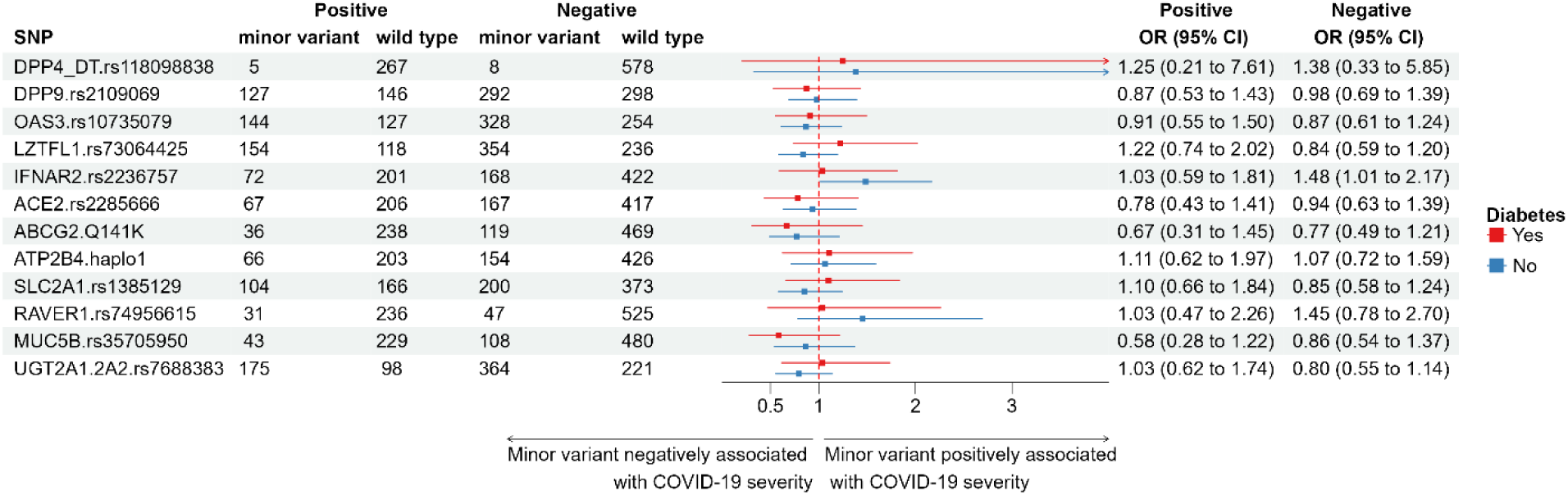
Associations between selected polymorphic genetic variations and disease severity in diabetic and nondiabetic COVID-19 patients. The figure provides the number of patients carrying the minor genetic variant (heterozygous+homozygous forms) and the wild-type variants for each parameter and shows a forest plot for the odds ratios (ORs) for associations of the selected parameters with disease severity. The forest plot indicates either lower or higher average OR values than 1 and the respective 95% confidence intervals severe, as compared to the mild COVID-19 cases.

### 4. Potential association of genetic polymorphisms with the anamnestic parameters of loss of smell and/or taste in patients with COVID-19

In the previous analysis, we found a significant association between lower disease severity and a reported loss of taste and/or smell (see Fig. 1). In the following, we analyzed the potential associations of the examined genetic polymorphisms with these anamnestic parameters. Since these anamnestic parameters are variable in clinical reports, we analyzed them separately.

In this case, we also analyzed the MAF values of the polymorphic variants of the examined genes in patients reporting or not reporting a loss of taste and/or smell, respectively. We also compared these MAF values to the values in the representative European population (see Supplementary Figure 3).

These data collectively indicate mostly similar effects as those for general hospitalized patients: COVID-19 patients with loss of taste or smell, carrying minor variants of *LZTFL1* and *RAVER1* (and probably also *ATP2B4, MUC5B* and *UGT2A1),* are more likely to have hospitalization-requiring COVID-19 disease than the general population. In contrast, patients who reported a loss of taste or smell and who carried minor variants of the *IFNAR2* gene (and probably also *OAS3)* were less likely to have COVID-19.

In the following, we analyzed the potential associations of genetic variants with the loss of taste and smell and the **severity** of COVID-19. Figure 5 shows the potential associations (ORs) of the examined genetic polymorphisms with COVID-19 disease severity in patients reporting a loss of taste (Figure 5 Panel A) and/or a loss of sense of smell. Again, due to the relatively smaller number of the respective patients, the confidence intervals on the graph for the potential associations of disease severity with these anamnestic reports are large in most cases for the examined genetic polymorphisms, and the differences in most cases do not reach statistical significance.

**Figure 5.**
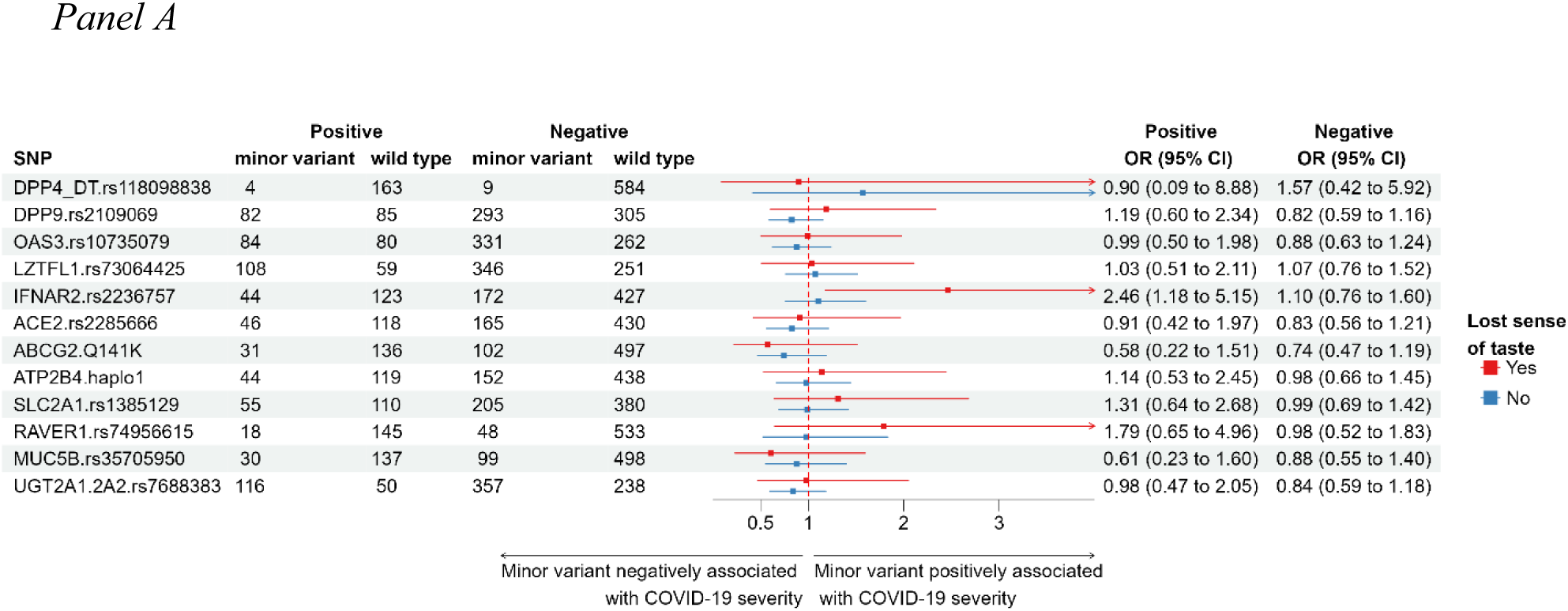
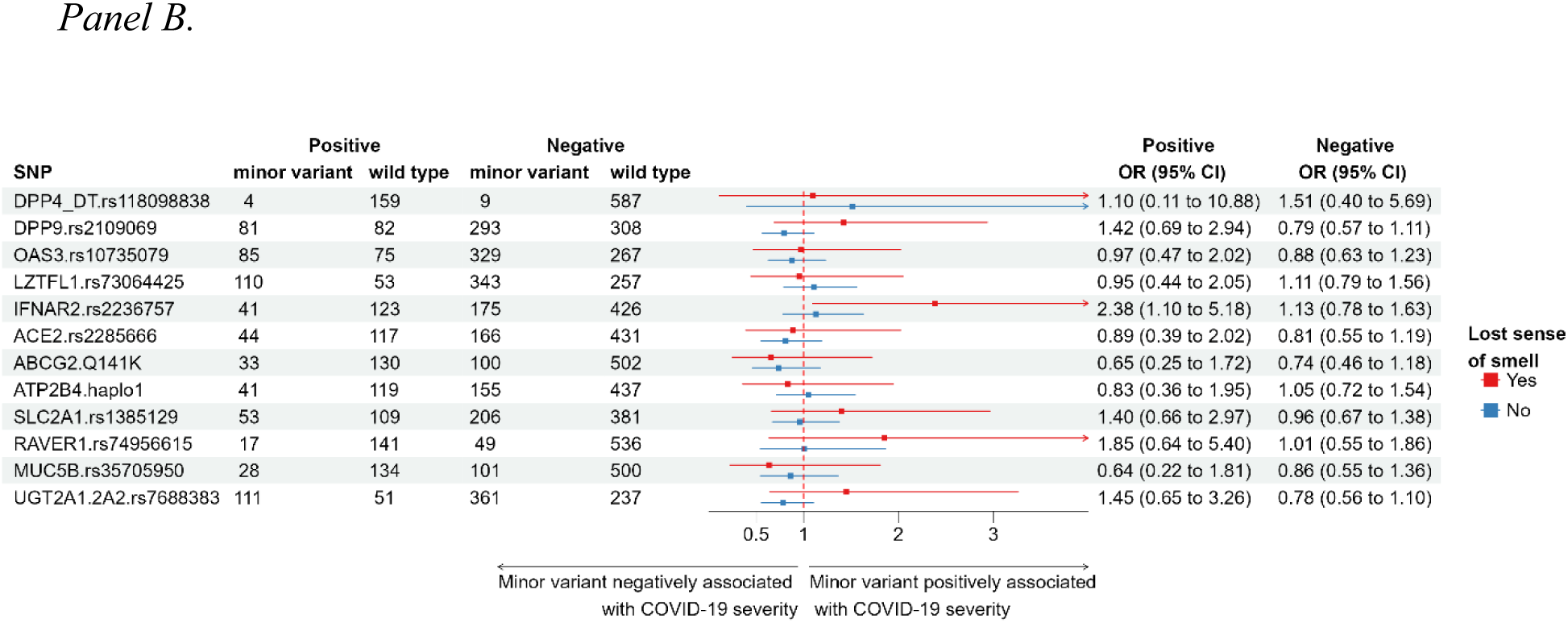
**Panel A.** Association of selected polymorphic genetic variations and disease severity in COVID-19 patients reporting loss of taste**. Panel B**. Association of selected polymorphic genetic variations and disease severity in COVID-19 patients reporting the loss of smell. The figures show the number of the respective patients carrying the minor genetic variant (heterozygous+homozygous forms) and the wild-type variants for each parameter and show forest plots for the odds ratios (ORs) for associations of the selected parameters with disease severity. The forest plot indicates either lower or higher average OR values than 1 and the respective 95% confidence intervals for severe, as compared to the mild COVID-19 cases.

Although the loss of these senses in the anamnesis generally correlates with a less severe COVID-19 disease (see Figure 2), the genetic polymorphisms have large variability regarding these parameters. Interestingly, the increased presence of the minor variant of IFNAR2 and probably also of RAVER1 and UGT2A1 in patients reporting a loss of taste or smell is still positively associated with severe disease.

In the Supplementary Materials (see Supplementary Figure 4), we provide additional information on the potential associations among the obtained anamnestic, clinical, and genetic data.

## Discussion

In the present work, we analyzed selected clinical and genetic data from 869 hospitalized COVID-19 patients in Hungary. Clinical data and blood samples were collected between 2020 and 2021, and the collection did not include effectively vaccinated patients. The dominant SARS-CoV-2 variants were Wuhan and the Delta variants. The detailed anamnestic and clinical laboratory data provided the opportunity to identify associations between these parameters, and for the genetic analyses, we selected the SNPs and the related haplotypes of 12 genes potentially relevant to the reaction to COVID-19 according to the GWAS-based literature.

Based on the data shown in the Results section and in the Supplementary Materials, we summarize the main points of this study:

1. Regarding the anamnestic parameters, preexisting diabetes had a major effect on the severity of COVID-19 at the clinic, while no such effects were observed for asthma, potentially because of the relatively low number of relevant patients. Interestingly, we found a significant association between the loss or altered sense of smell or taste and a less severe clinical form of COVID-19. Related to these findings, among patients with diabetes, loss of smell or taste occurred less frequently in diabetic than in nondiabetic patients (a detailed analysis of the clinical data is provided in a follow-up paper).
2. When looking for potential associations between the genetic variants in the 12 genes studied, we performed several types of analyses. First, we compared the minor allele frequency values in clinically treated COVID-19 patients to the MAF values in the general European population. We performed a similar analysis in patients with different disease severities or preexisting conditions. The Hungarian population in all aspects closely reflects the European genetic SNP patterns (see refs [41–43]), thus the MAF datasets for European population, provided by the 1000 Genome (phase3 release V3+) and the ALFA (Release Version: 20230706150541) databases, are most reliable for assessing population-based genetic differences. Although these databases, in some cases, provide variable data (see Supplementary Table 1 and Supplementary Figure 1), they are still more relevant for such an analysis than a few hundred control MAF values obtained locally.

Another way of analyzing the associations of genetic factors with disease conditions was to directly compare the presence of minor allele variants between the categories of different disease severities or key anamnestic parameters. According to these combined analyses, the following conclusions can be drawn:

a. Most of the gene polymorphisms analyzed had important associations with COVID-19. Regarding the hospitalized patients, a relatively higher mean level of the *LZTFL1* minor variant (higher MAF COVID/MAF EU population) significantly correlated with greater COVID-19 disease susceptibility. A similar tendency was observed for *ATP2B4* haplotype 1, *RAVER1, MUC5B,* and *UGT2A1* variants, but these differences were significant only compared to the ALFA population values. These data are in accordance with those in the literature (see refs [10–14, 17–24, 27]).

In contrast to some of the data in the literature, we found that a relatively lower mean level of the minor variant of the *IFNAR2* gene (lower MAF COVID/MAF EU population) in hospitalized patients correlated with lower COVID-19 disease susceptibility, and this was also observed in diabetic patients. Recent clinical and genetic data also question the previously indicated role of this variant in disease severity [28, 44, 45].

The SNPs that did not show any significant associations with COVID-19-related hospitalization in our analysis were *DPP4-DT* (probably because of the low MAF values and the relatively small population), *DPP9*, *ABCG2*, and *SLC2A1*.

b. When examining the associations of genetic variations with the severity of COVID-19 disease, in accordance with the data in the literature, we observed that a higher mean level of the *LZTFL1* minor variant correlated with greater disease severity, and a similar tendency was observed in the cases of the *RAVER1, MUC5B,* and *UGT2A1* variants, but these trends were significant only compared to the ALFA population values. In this analysis, we found again that a relatively lower mean level of the minor variant of the *IFNAR2* gene (lower MAF COVID/MAF EU population) in hospitalized patients correlated with lower COVID-19 severity.

c. In COVID-19 patients who also had diabetes, the *LZTFL1* and *IFNAR2* minor variants (MAF values) showed a similar inverse association. Interestingly, in diabetic COVID-19 patients, the mean MAF for *ABCG2* was lower (correlating with reduced disease susceptibility), while the opposite trend was observed for *RAVER1*. In COVID-19 patients with diabetes, we also found that a lower mean level of the minor variant of the IFNAR2 gene correlated with a lower susceptibility to COVID-19. An interesting potential genetic association was observed between the reduced-function ABCG2 transporter variant and disease severity, as well as in diabetic patients. In these cases, the presence of this rather common genetic variant seems to be associated with a protective effect, although these results were not statistically significant. Further exploration of this genetic association should be performed in a larger patient population and potentially in patients with long-term COVID-19. In our additional studies, in patients reporting a loss of smell or taste, the examined genetic associations showed a pattern similar to that in the general hospitalized COVID-19 population.

When disease-related SNPs are identified through GWAS, these variables often have only small effects on the respective ORs and may be lost in the background noise. By using high-level statistical thresholds, only genetic variants with strong statistical evidence can be regarded as significant; thus, variants with smaller effects are filtered out. Especially in infectious diseases with variable tissue pathologies, multiple genetic variants with small individual effects may contribute to the overall risk. Therefore, our current study, although it focused only on a limited number of potentially relevant genetic variants in a relatively small number of patients, may allow a focused, targeted analysis of clinical and genetic data with potential relevance to the COVID-19 clinic.

The data in this study may help to decipher the association between selected genetic variants and the incidence of COVID-19. The data were obtained for people infected with the Wuhan and Delta variants; thus, they may not be relevant to the clinical effects of the newly emerging SARS-CoV-2 variants, e.g., the Omicron variants. Nevertheless, the basic clinical problems in severe cases of current diseases involve the same cellular virus receptors and the overreaction of the immune system (see [46, 47]). Most of the genetic variations studied here are related to virus receptors, the virus-activated immune system, or general metabolic regulators.

A newly emerging question is the potential genetic background of the rapidly increasing number of long-term or post-COVID-19 patients with currently ill-defined clinical symptoms (see [48–54]). When extended to the current COVID-19 situation and using larger patient numbers, our approach may provide a personalized tool for assessing the expected course, severity, and long-term, chronic effects in COVID-19 patients.

## Methods

### Clinical samples

Detailed clinical data were entered into this database by the participating clinicians at the Korányi Clinic (led by Judit Moldvay) and the University of Pécs, with the leadership of Péter Hegyi. The patients were informed about the research project, and written consent was obtained to participate in this study. All methods were performed in accordance with the relevant guidelines and regulations. Ethical permission from ETT TUKEB, NNK 24004-7/2021/EihO, and 20800-6/2020/EÜIG was obtained to perform these noninvasive molecular genetic studies. Clinical data and blood samples were collected between 2020 and 2021; at that time, no vaccination was accessible, and the collection did not include effectively vaccinated patients.

The anonymized data presented in this study, as well as those of the genetic analyses, are available in the data repository upon request.

### Genetic analysis

In this work, we studied the prevalence of specific human SNPs and their potential alteration frequency in (n=869) COVID-19 patients. The SNPs studied here are summarized in Supplementary Table 1.

We prepared genomic DNA from blood samples obtained from COVID-19 patients with variable clinical courses (see above). Blood samples (1 mL) were collected in EDTA tubes during routine laboratory testing (without any additional burden to the patients). Genomic DNA was purified from 300 μL of EDTA-anticoagulated blood samples with a Puregene Blood Kit (Qiagen). TaqMan-based qPCRs for SNPs (for details, see Suppl. Table 2) were performed in a StepOnePlus device (Applied Biosystems) with premade assay mixes and a master mix (cat. 4371353) from Thermo Fisher. The results of the molecular genetic studies and the clinical and laboratory data were collected in an anonymized database and subjected to detailed statistical analysis.

### Statistical analysis

The European minor allele frequencies (ALFA and 1000G) were collected from the NCBI dbSNP database (downloading date: 10/1/2024, https://www.ncbi.nlm.nih.gov/snp/). Statistical analysis was conducted with GraphPad Prism 8.0.1.

The odds ratios (ORs) and 95% confidence intervals (CIs) for the associations between severity and the SNPs and/or comorbidities were calculated by logistic regression (R Studio, version number: 4.1.3). The comparisons of the allele frequencies with the European population values were analyzed by using Fisher’s exact test (p<0.05), and the ORs with confidence intervals were analyzed by the Baptista-Pike test (Prism 8.0.1, GraphPad). The number of patients (n) involved in each analysis is indicated in the respective tables. Additional statistical analyses and methods are described in the Supplementary Materials.

## Supporting information

Supplemental Materials Mozner et al

## Acknowledgments

We gratefully thank Dr. Andrea Harnos and Márk Bérchegyi for their help in the statistical analyses. We are thankful to Dr. Szilárd Tóth for his help with the automated pipetting preparation of the qPCR plates. This research was supported by the Post-COVID Research Program of the Hungarian Academy of Sciences (PC2022-7/2022, Balazs Sarkadi.); project no. KDP-1017403, O.M., has been implemented with support provided by the Ministry of Culture and Innovation of Hungary from the National Research, Development and Innovation Fund, financed under the KDP-2020 funding scheme. Project K-128011 (Gy. V.) has been implemented with the support provided by the Ministry of Culture and Innovation from the National Research, Development and Innovation Fund. This research was also supported by NKFIH OTKA grants (K131996 and K147265) to PH and the University of Pécs Medical School Research Fund (300909) to AS.

## Institutional Review Board Statement

This project was carried out with the ethical permission of 24004-7/2021/EihO, issued on 7 May 2021, by the NNK Hungary - ETT TUKEB and 20800-6/2020/EÜIG, issued on 5 May 2020, by the NNK Hungary - ETT TUKEB.

## Informed Consent Statement

Informed consent was obtained from all the subjects involved in the study.

## Authors’ contributions

O.M., E.Sz. and A.K. designed and performed the experiments and analyzed the data; E.Sz. and A.K. performed the bioinformatic analysis; V.V., A.Sz., and Á.J. provided and reviewed the clinical data. O.M., E.Sz., A.K., Gy.V., P.H. and B.S. contributed to the writing and critical revision of the manuscript. All the authors have read and approved the final version.

## Data Availability

The datasets generated and analyzed during the current study are available from the corresponding authors on reasonable request. The anonymized data for the anamnestic and clinical laboratory data, as well as those of the genetic analyses are available in the data repository upon request.

