## Supplemental Materials Mozner et al for "Potential associations of selected polymorphic genetic variants with COVID-19 disease susceptibility and severity"

**Supplementary Materials for Mózner et al: Potential correlation of selected polymorphic genetic variants with COVID-19 disease susceptibility and severity**

**Supplementary table 1: SNPs analyzed and their minor allele frequency (MAF) in the European population.** MAF (Eur) from the dbSNP databases (see **https://www.ncbi.nlm.nih.gov/snp/)**

| SNP | MAF (Eur) (ALFA/1000G) | Related gene(s) | COVID-19 or other disease association |
|---|---|---|---|
| rs118098838 (chr 2) | 0.0050/ 0.0099 | *DPP4-DT*, NRP1 and NRP2 | Severe COVID-19 symptoms |
| rs2109069 (chr 12) | 0.3088/ 0.3211 | *DPP9* | Severe COVID-19 symptoms |
| rs10735079 (chr 12) | 0.3662/ 0.3638 | *OAS3* | More/less severe COVID-19 symptoms |
| rs73064425 (chr 3) | 0.0820/ 0.0795 | *LZTFL1* | Severe COVID-19 symptoms |
| rs2236757 (chr 21) | 0.2941/ 0.2942 | *IFNAR2* | Severe COVID-19 symptoms |
| rs2285666 (chr X) | 0.2038/ 0.2350 | *ACE2* | Affects ACE2 protein expression |
| rs2231142 (chr 4) | 0.1032/ 0.0944 | *ABCG2* | Decreased ABCG2 protein expression, gout susceptibility |
| rs1541252 (chr 1) | 0.1082/ 0.1024 | *ATP2B4* | Regulation of PMCA4b expression |
| rs1385129 (chr 1) | 0.2120/ 0.2187 | *SLC2A1* | Poor CD4+ T cell recovery in antiretroviral-treated individuals |
| rs74956615 (chr 19) | 0.0211/ 0.0298 | *RAVER1* | Associated with critical illness in COVID-19 |
| rs35705950 (chr 11) | 0.0352/ 0.1074 | *MUC5B* | Associated with mucin accumulation in bronchi and alveolar ducts in COVID-19 |
| rs7688383 (chr 4) | 0.3664/ 0.3857 | *UGT2A1* | Associated with loss of smell or taste in COVID-19 |

#### Supplementary table 2: SNPs and DNA sequences of the probes used in qPCR experiments

| SNPs | Order ID | **Context Sequence [VIC/FAM]** |
|---|---|---|
| rs118098838 | C_150493403_10 | CCTTTATCATATAGATACAATTTTC[G/T]CTTAATAATAGCATGCTCCTGCAAA |
| rs2109069 | C__11517118_10 | TCACCCAGAGAGGAAGGGGAGTGGA[G/A]CCCCAGTCTCTTGGAGCCCAAAACC |
| rs10735079 | C__31831768_10 | TGGCTAGCAGTAGGGGCCTGGGGAC[A/G]AAACCAGAATTCTGCAAAATCTTGT |
| rs2236757 | C__11354003_30 | CAAATCCCAAAAGAGATTAAGGCCT[A/G]CCTCTAAATGAAATTCTCAGTCTTA |
| rs73064425 | C__98755833_10 | CTCATTTTTAAATGACAAAAATTAA[C/T]GAATGAATGAAAGTGGATCTTTCAC |
| rs2285666 | C___2551626_1_ | ATAATCACTACTAAAAATTAGTAGC[C/T]TACCTGGTTCAAGTAATAAGCATTC |
| rs1541252 | C____360682_10 | TCCTCTTCCTCCTCCTGACGTCTAC[C/T]ACTACAGTTGCTGGTTGTTGCTAAG |
| rs1385129 | C___1166185_1_ | GGGAGCCAAGCACTGCTCCTCCCAC[A/G]GCCAGCATGAGGCGACCCGTCAGCT |
| rs2231142 | C__15854163_70 | GCAAGCCGAAGAGCTGCTGAGAACT[G/T]TAAGTTTTCTCTCACCGTCAGAGTG |
| rs74956615 | C__27854626_20 | TAGAAAAGGAAACAGAAGTCAGTTG[T/A]CAAAGTTAAAAAAAAAGGAGACAGT |
| rs35705950 | C___1582254_20 | CCTTCCTTTATCTTCTGTTTTCAGC[G/T]CCTTCAACTGTGAAGAGGTGAACTC |
| rs7688383 | C___2951323_10 | TTTATTGCAGTACTGTTCCCAATAG[C/T]CAAGATATGGAATCAACCTATATGT |

**Supplementary table 3: Statistical analysis of the clinical parameters, the related MAF values and the H-W distributions for each genetic variant and Khi$^2$ probes for each group examined.** The left tables show the number of each examined group. Right tables show the calculated minor allele frequencies (MAF) and Hardy-Weinberg Equilibrium (HWE) every groups. The Khi$^2$ test were calculated the number of wild type (wt) and minor variants (MV) in pair groups (male/female, diabetes yes/no, diabetes/T1DM, diabetes/T2DM, asthma yes/no, loss of taste yes/no, loss of smell yes/no, severity mild/severe, death yes/no). diab: diabetes, T1DM: type 1 diabetes, T2DM: type 2 diabetes, mod: moderate severity, ser: severe severity, crit: critical severity, mild+mod is a group of mild in the article, ser+crit is a group of severe in the article.

### DPP4-DT

| n | wt | hetero | homo | wt | MV |
|---|---|---|---|---|---|
| all | 846 | 13 | 0 | 846 | 13 |
| male | 452 | 4 | 0 | 452 | 4 |
| female | 394 | 9 | 0 | 394 | 9 |
| diab no | 578 | 8 | 0 | 578 | 8 |
| diab yes | 267 | 5 | 0 | 267 | 5 |
| T1DM | 16 | 1 | 0 | 16 | 1 |
| T2DM | 247 | 4 | 0 | 247 | 4 |
| asthma no | 793 | 12 | 0 | 793 | 12 |
| asthma yes | 52 | 1 | 0 | 52 | 1 |
| taste no | 584 | 9 | 0 | 584 | 9 |
| taste yes | 163 | 4 | 0 | 163 | 4 |
| smell no | 587 | 9 | 0 | 587 | 9 |
| smell yes | 159 | 4 | 0 | 159 | 4 |
| mod | 386 | 7 | 0 | 386 | 7 |
| ser | 139 | 2 | 0 | 139 | 2 |
| mild | 192 | 1 | 0 | 192 | 1 |
| crit | 129 | 3 | 0 | 129 | 3 |
| mild+mod | 578 | 8 | 0 | 578 | 8 |
| ser+crit | 268 | 5 | 0 | 268 | 5 |
| death no | 710 | 11 | 0 | 710 | 11 |
| death yes | 136 | 2 | 0 | 136 | 2 |

| | MAF | HWE | Khi$^2$ |
|---|---|---|---|
| all | 0.0076 | 0.9755 | |
| male | 0.0044 | 0.9956 | 0.1042 |
| female | 0.0112 | 0.9749 | |
| diab no | 0.0068 | 0.9863 | 0.5976 |
| diab yes | 0.0092 | 0.9885 | |
| T1DM | 0.0294 | 0.9925 | 0.1300 |
| T2DM | 0.0080 | 0.9920 | 0.7989 |
| asthma no | 0.0075 | 0.9777 | 0.8191 |
| asthma yes | 0.0094 | 0.9976 | |
| taste no | 0.0076 | 0.9829 | 0.4398 |
| taste yes | 0.0120 | 0.9880 | |
| smell no | 0.0076 | 0.9830 | 0.4105 |
| smell yes | 0.0123 | 0.9877 | |
| mod | 0.0089 | 0.9844 | |
| ser | 0.0071 | 0.9964 | |
| mild | 0.0026 | 0.9994 | |
| crit | 0.0114 | 0.9914 | |
| mild+mod | 0.0068 | 0.9863 | 0.6022 |
| ser+crit | 0.0092 | 0.9885 | |
| death no | 0.0076 | 0.9791 | 0.9463 |
| death yes | 0.0072 | 0.9964 | |

### DPP9

| n | wt | hetero | homo | wt | MV |
|---|---|---|---|---|---|
| all | 444 | 328 | 92 | 444 | 420 |
| male | 235 | 181 | 47 | 235 | 228 |
| female | 209 | 147 | 45 | 209 | 192 |
| diab no | 298 | 230 | 62 | 298 | 292 |
| diab yes | 146 | 97 | 30 | 146 | 127 |
| T1DM | 12 | 5 | 1 | 12 | 6 |
| T2DM | 131 | 92 | 28 | 131 | 120 |
| asthma no | 415 | 307 | 89 | 415 | 396 |
| asthma yes | 29 | 20 | 3 | 29 | 23 |
| taste no | 305 | 231 | 62 | 305 | 293 |
| taste yes | 85 | 64 | 18 | 85 | 82 |
| smell no | 308 | 230 | 63 | 308 | 293 |
| smell yes | 82 | 64 | 17 | 82 | 81 |
| mod | 193 | 157 | 44 | 193 | 201 |
| ser | 75 | 52 | 13 | 75 | 65 |
| mild | 106 | 65 | 23 | 106 | 88 |
| crit | 70 | 54 | 12 | 70 | 66 |
| mild+mod | 299 | 222 | 67 | 299 | 289 |
| ser+crit | 145 | 106 | 25 | 145 | 131 |
| death no | 368 | 276 | 79 | 368 | 355 |
| death yes | 76 | 52 | 13 | 76 | 65 |

| | MAF | HWE | $Khi^2$ |
|---|---|---|---|
| all | 0.2963 | 0.1856 | |
| male | 0.2970 | 0.6310 | 0.6892 |
| female | 0.2955 | 0.2517 | |
| diab no | 0.3000 | 0.4756 | 0.4167 |
| diab yes | 0.2875 | 0.3167 | |
| T1DM | 0.1944 | 0.9496 | 0.1767 |
| T2DM | 0.2948 | 0.4283 | 0.6551 |
| asthma no | 0.2990 | 0.1573 | 0.5202 |
| asthma yes | 0.2500 | 0.9913 | |
| taste no | 0.2968 | 0.4442 | 0.9808 |
| taste yes | 0.2994 | 0.7381 | |
| smell no | 0.2962 | 0.3739 | 0.8312 |
| smell yes | 0.3006 | 0.8396 | |
| mod | 0.3109 | 0.6228 | |
| ser | 0.2786 | 0.8226 | |
| mild | 0.2861 | 0.2277 | |
| crit | 0.2868 | 0.9715 | |
| mild+mod | 0.3027 | 0.2039 | 0.6439 |
| ser+crit | 0.2826 | 0.8283 | |
| death no | 0.3001 | 0.2313 | 0.5142 |
| death yes | 0.2766 | 0.8109 | |

### OAS3

| n | wt | hetero | homo | wt | MV |
|---|---|---|---|---|---|
| all | 382 | 386 | 86 | 382 | 472 |
| male | 201 | 211 | 42 | 201 | 253 |
| female | 181 | 175 | 44 | 181 | 219 |
| diab no | 254 | 267 | 61 | 254 | 328 |
| diab yes | 127 | 119 | 25 | 127 | 144 |
| T1DM | 9 | 8 | 0 | 9 | 8 |
| T2DM | 115 | 110 | 25 | 115 | 135 |
| asthma no | 366 | 352 | 84 | 366 | 436 |
| asthma yes | 15 | 34 | 2 | 15 | 36 |
| taste no | 262 | 271 | 60 | 262 | 331 |
| taste yes | 80 | 69 | 15 | 80 | 84 |
| smell no | 267 | 270 | 59 | 267 | 329 |
| smell yes | 75 | 70 | 15 | 75 | 85 |
| mod | 172 | 177 | 38 | 172 | 215 |
| ser | 68 | 58 | 11 | 68 | 69 |
| mild | 82 | 88 | 24 | 82 | 112 |
| crit | 60 | 63 | 13 | 60 | 76 |
| mild+mod | 254 | 265 | 62 | 254 | 327 |
| ser+crit | 128 | 121 | 24 | 128 | 145 |
| death no | 319 | 320 | 74 | 319 | 394 |
| death yes | 63 | 66 | 12 | 63 | 78 |

| | MAF | HWE | $Khi^2$ |
|---|---|---|---|
| all | 0.3267 | 0.8507 | |
| male | 0.3249 | 0.6644 | 0.7745 |
| female | 0.3288 | 0.9924 | |
| diab no | 0.3342 | 0.8694 | 0.3783 |
| diab yes | 0.3118 | 0.9640 | |
| T1DM | 0.2353 | 0.5382 | 0.4464 |
| T2DM | 0.3200 | 0.9924 | 0.5303 |
| asthma no | 0.3242 | 0.9995 | 0.0238 |
| asthma yes | 0.3725 | 0.0686 | |
| taste no | 0.3297 | 0.8413 | 0.2950 |
| taste yes | 0.3018 | 0.9999 | |
| smell no | 0.3255 | 0.8598 | 0.6394 |
| smell yes | 0.3125 | 0.9868 | |
| mod | 0.3269 | 0.8595 | |
| ser | 0.2920 | 0.9803 | |
| mild | 0.3505 | 0.9993 | |
| crit | 0.3272 | 0.9104 | |
| mild+mod | 0.3348 | 0.9189 | 0.3851 |
| ser+crit | 0.3095 | 0.9100 | |
| death no | 0.3282 | 0.9449 | 0.9896 |
| death yes | 0.3191 | 0.8064 | |

### LZTFL1

| n | wt | hetero | homo | wt | MV |
|---|---|---|---|---|---|
| all | 355 | 399 | 109 | 355 | 508 |
| male | 191 | 209 | 61 | 191 | 270 |
| female | 164 | 190 | 48 | 164 | 238 |
| diab no | 236 | 277 | 77 | 236 | 354 |
| diab yes | 118 | 122 | 32 | 118 | 154 |
| T1DM | 8 | 5 | 5 | 8 | 10 |
| T2DM | 108 | 116 | 26 | 108 | 142 |
| asthma no | 335 | 373 | 103 | 335 | 476 |
| asthma yes | 19 | 26 | 6 | 19 | 32 |
| taste no | 251 | 273 | 73 | 251 | 346 |
| taste yes | 59 | 83 | 25 | 59 | 108 |
| smell no | 257 | 271 | 72 | 257 | 343 |
| smell yes | 53 | 84 | 26 | 53 | 110 |
| mod | 158 | 183 | 52 | 158 | 235 |
| ser | 57 | 61 | 23 | 57 | 84 |
| mild | 81 | 91 | 22 | 81 | 113 |
| crit | 59 | 64 | 12 | 59 | 76 |
| mild+mod | 239 | 274 | 74 | 239 | 348 |
| ser+crit | 116 | 125 | 35 | 116 | 160 |
| death no | 291 | 337 | 95 | 291 | 432 |
| death yes | 64 | 62 | 14 | 64 | 76 |

| | MAF | HWE | $Khi^2$ |
|---|---|---|---|
| all | 0.3575 | 0.9910 | |
| male | 0.3590 | 0.9747 | 0.8499 |
| female | 0.3557 | 0.9067 | |
| diab no | 0.3653 | 0.9771 | 0.3482 |
| diab yes | 0.3419 | 0.9993 | |
| T1DM | 0.4167 | 0.4265 | 0.7047 |
| T2DM | 0.3360 | 0.9045 | 0.3885 |
| asthma no | 0.3570 | 0.9993 | 0.5683 |
| asthma yes | 0.3725 | 0.8994 | |
| taste no | 0.3509 | 0.9978 | 0.1183 |
| taste yes | 0.3982 | 0.9443 | |
| smell no | 0.3458 | 0.9995 | 0.0174 |
| smell yes | 0.4172 | 0.8640 | |
| mod | 0.3651 | 0.9981 | |
| ser | 0.3794 | 0.7934 | |
| mild | 0.3479 | 0.9458 | |
| crit | 0.3259 | 0.8060 | |
| mild+mod | 0.3595 | 0.9730 | 0.7146 |
| ser+crit | 0.3533 | 0.9946 | |
| death no | 0.3645 | 0.9931 | 0.2290 |
| death yes | 0.3214 | 0.9919 | |

### IFNAR2

| n | wt | hetero | homo | wt | MV |
|---|---|---|---|---|---|
| all | 624 | 213 | 27 | 624 | 240 |
| male | 335 | 114 | 15 | 335 | 129 |
| female | 289 | 99 | 12 | 289 | 111 |
| diab no | 422 | 147 | 21 | 422 | 168 |
| diab yes | 201 | 66 | 6 | 201 | 72 |
| T1DM | 12 | 6 | 0 | 12 | 6 |
| T2DM | 186 | 59 | 6 | 186 | 65 |
| asthma no | 587 | 200 | 23 | 587 | 223 |
| asthma yes | 36 | 13 | 4 | 36 | 17 |
| taste no | 427 | 149 | 23 | 427 | 172 |
| taste yes | 123 | 40 | 4 | 123 | 44 |
| smell no | 426 | 151 | 24 | 426 | 175 |
| smell yes | 123 | 38 | 3 | 123 | 41 |
| mod | 283 | 104 | 8 | 283 | 112 |
| ser | 99 | 33 | 8 | 99 | 41 |
| mild | 153 | 37 | 4 | 153 | 41 |
| crit | 89 | 39 | 7 | 89 | 46 |
| mild+mod | 436 | 141 | 12 | 436 | 153 |
| ser+crit | 188 | 72 | 15 | 188 | 87 |
| death no | 526 | 175 | 23 | 526 | 198 |
| death yes | 98 | 38 | 4 | 98 | 42 |

| | MAF | HWE | $Khi^2$ |
|---|---|---|---|
| all | 0.1545 | 0.5338 | |
| male | 0.1552 | 0.6602 | 0.9865 |
| female | 0.1538 | 0.8023 | |
| diab no | 0.1602 | 0.4861 | 0.5218 |
| diab yes | 0.1429 | 0.9891 | |
| T1DM | 0.1667 | 0.7404 | 0.6532 |
| T2DM | 0.1414 | 0.9410 | 0.4446 |
| asthma no | 0.1519 | 0.7236 | 0.4744 |
| asthma yes | 0.1981 | 0.5619 | |
| taste no | 0.1628 | 0.3644 | 0.5477 |
| taste yes | 0.1437 | 0.9719 | |
| smell no | 0.1656 | 0.3359 | 0.2991 |
| smell yes | 0.1341 | 0.9997 | |
| mod | 0.1519 | 0.9518 | |
| ser | 0.1750 | 0.3784 | |
| mild | 0.1160 | 0.8189 | |
| crit | 0.1963 | 0.8018 | |
| mild+mod | 0.1401 | 0.9943 | 0.0836 |
| ser+crit | 0.1855 | 0.3446 | |
| death no | 0.1526 | 0.4976 | 0.5213 |
| death yes | 0.1643 | 0.9955 | |

### ACE2

| n | wt | hetero | homo | wt | MV |
|---|---|---|---|---|---|
| all | 624 | 121 | 113 | 624 | 234 |
| male | 361 | 3 | 92 | 361 | 95 |
| female | 263 | 118 | 21 | 263 | 139 |
| diab no | 417 | 84 | 83 | 417 | 167 |
| diab yes | 206 | 37 | 30 | 206 | 67 |
| T1DM | 15 | 3 | 0 | 15 | 3 |
| T2DM | 187 | 34 | 30 | 187 | 64 |
| asthma no | 589 | 108 | 108 | 589 | 216 |
| asthma yes | 34 | 13 | 5 | 34 | 18 |
| taste no | 430 | 85 | 80 | 430 | 165 |
| taste yes | 118 | 24 | 22 | 118 | 46 |
| smell no | 431 | 87 | 79 | 431 | 166 |
| smell yes | 117 | 21 | 23 | 117 | 44 |
| mod | 274 | 56 | 60 | 274 | 116 |
| ser | 103 | 25 | 12 | 103 | 37 |
| mild | 146 | 26 | 22 | 146 | 48 |
| crit | 101 | 14 | 19 | 101 | 33 |
| mild+mod | 420 | 82 | 82 | 420 | 164 |
| ser+crit | 204 | 39 | 31 | 204 | 70 |
| death no | 518 | 107 | 94 | 518 | 201 |
| death yes | 106 | 14 | 19 | 106 | 33 |

| | MAF | HWE | $Khi^2$ |
|---|---|---|---|
| all | 0.2022 | 0.0000 | |
| male | 0.2050 | 0.0000 | 0.0000 |
| female | 0.1990 | 0.5578 | |
| diab no | 0.2140 | 0.0000 | 0.2146 |
| diab yes | 0.1777 | 0.0000 | |
| T1DM | 0.0833 | 0.9341 | 0.2681 |
| T2DM | 0.1873 | 0.0000 | 0.3589 |
| asthma no | 0.2012 | 0.0000 | 0.2221 |
| asthma yes | 0.2212 | 0.4332 | |
| taste no | 0.2059 | 0.0000 | 0.9359 |
| taste yes | 0.2073 | 0.0000 | |
| smell no | 0.2052 | 0.0000 | 0.9046 |
| smell yes | 0.2081 | 0.0000 | |
| mod | 0.2256 | 0.0000 | |
| ser | 0.1750 | 0.0225 | |
| mild | 0.1804 | 0.0000 | |
| crit | 0.1940 | 0.0000 | |
| mild+mod | 0.2106 | 0.0000 | 0.4370 |
| ser+crit | 0.1843 | 0.0000 | |
| death no | 0.2051 | 0.0000 | 0.3071 |
| death yes | 0.1871 | 0.0000 | |

### ABCG2

| n | wt | hetero | homo | wt | MV |
|---|---|---|---|---|---|
| all | 708 | 144 | 11 | 708 | 155 |
| male | 372 | 81 | 8 | 372 | 89 |
| female | 336 | 63 | 3 | 336 | 66 |
| diab no | 469 | 110 | 9 | 469 | 119 |
| diab yes | 238 | 34 | 2 | 238 | 36 |
| T1DM | 15 | 3 | 0 | 15 | 3 |
| T2DM | 219 | 31 | 2 | 219 | 33 |
| asthma no | 663 | 136 | 11 | 663 | 147 |
| asthma yes | 44 | 8 | 0 | 44 | 8 |
| taste no | 497 | 96 | 6 | 497 | 102 |
| taste yes | 136 | 28 | 3 | 136 | 31 |
| smell no | 502 | 93 | 7 | 502 | 100 |
| smell yes | 130 | 31 | 2 | 130 | 33 |
| mod | 323 | 66 | 6 | 323 | 72 |
| ser | 127 | 13 | 1 | 127 | 14 |
| mild | 151 | 38 | 4 | 151 | 42 |
| crit | 107 | 27 | 0 | 107 | 27 |
| mild+mod | 474 | 104 | 10 | 474 | 114 |
| ser+crit | 234 | 40 | 1 | 234 | 41 |
| death no | 596 | 116 | 11 | 596 | 127 |
| death yes | 112 | 28 | 0 | 112 | 28 |

| | MAF | HWE | Khi$^2$ |
|---|---|---|---|
| all | 0.0962 | 0.7371 | |
| male | 0.1052 | 0.6530 | 0.2702 |
| female | 0.0858 | 0.9998 | |
| diab no | 0.1088 | 0.8448 | 0.0115 |
| diab yes | 0.0693 | 0.9193 | |
| T1DM | 0.0833 | 0.9341 | 0.7097 |
| T2DM | 0.0694 | 0.8906 | 0.0137 |
| asthma no | 0.0975 | 0.6892 | 0.6149 |
| asthma yes | 0.0769 | 0.8465 | |
| taste no | 0.0902 | 0.9298 | 0.6434 |
| taste yes | 0.1018 | 0.7958 | |
| smell no | 0.0889 | 0.7634 | 0.2775 |
| smell yes | 0.1074 | 0.9976 | |
| mod | 0.0987 | 0.7365 | |
| ser | 0.0532 | 0.8552 | |
| mild | 0.1192 | 0.8516 | |
| crit | 0.1007 | 0.4693 | |
| mild+mod | 0.1054 | 0.6185 | 0.1102 |
| ser+crit | 0.0764 | 0.9235 | |
| death no | 0.0954 | 0.4847 | 0.4922 |
| death yes | 0.1000 | 0.4593 | |

### ATP2B4

| n | wt | hetero | homo | wt | MV |
|---|---|---|---|---|---|
| all | 630 | 208 | 12 | 630 | 220 |
| male | 333 | 115 | 7 | 333 | 122 |
| female | 297 | 93 | 5 | 297 | 98 |
| diab no | 426 | 143 | 11 | 426 | 154 |
| diab yes | 203 | 65 | 1 | 203 | 66 |
| T1DM | 14 | 4 | 0 | 14 | 4 |
| T2DM | 185 | 61 | 1 | 185 | 62 |
| asthma no | 589 | 196 | 11 | 589 | 207 |
| asthma yes | 40 | 12 | 1 | 40 | 13 |
| taste no | 438 | 143 | 9 | 438 | 152 |
| taste yes | 119 | 42 | 2 | 119 | 44 |
| smell no | 437 | 146 | 9 | 437 | 155 |
| smell yes | 119 | 39 | 2 | 119 | 41 |
| mod | 288 | 90 | 7 | 288 | 97 |
| ser | 100 | 38 | 1 | 100 | 39 |
| mild | 143 | 48 | 2 | 143 | 50 |
| crit | 99 | 32 | 2 | 99 | 34 |
| mild+mod | 431 | 138 | 9 | 431 | 147 |
| ser+crit | 199 | 70 | 3 | 199 | 73 |
| death no | 530 | 171 | 10 | 530 | 181 |
| death yes | 100 | 37 | 2 | 100 | 39 |

| | MAF | HWE | $Khi^2$ |
|---|---|---|---|
| all | 0.1365 | 0.7108 | |
| male | 0.1418 | 0.8299 | 0.5060 |
| female | 0.1304 | 0.8520 | |
| diab no | 0.1422 | 0.9841 | 0.5327 |
| diab yes | 0.1245 | 0.3174 | |
| T1DM | 0.1111 | 0.8824 | 0.6816 |
| T2DM | 0.1275 | 0.3408 | 0.6639 |
| asthma no | 0.1369 | 0.6811 | 0.8122 |
| asthma yes | 0.1321 | 0.9980 | |
| taste no | 0.1364 | 0.8793 | 0.7512 |
| taste yes | 0.1411 | 0.8277 | |
| smell no | 0.1385 | 0.8366 | 0.8867 |
| smell yes | 0.1344 | 0.9019 | |
| mod | 0.1351 | 1.0000 | |
| ser | 0.1439 | 0.5707 | |
| mild | 0.1347 | 0.7729 | |
| crit | 0.1353 | 0.9724 | |
| mild+mod | 0.1349 | 0.9249 | 0.6625 |
| ser+crit | 0.1397 | 0.6662 | |
| death no | 0.1343 | 0.7974 | 0.5220 |
| death yes | 0.1475 | 0.8729 | |

### SLC2A1

| n | wt | hetero | homo | wt | MV |
|---|---|---|---|---|---|
| all | 540 | 265 | 39 | 540 | 304 |
| male | 282 | 149 | 20 | 282 | 169 |
| female | 258 | 116 | 19 | 258 | 135 |
| diab no | 373 | 176 | 24 | 373 | 200 |
| diab yes | 166 | 89 | 15 | 166 | 104 |
| T1DM | 9 | 7 | 1 | 9 | 8 |
| T2DM | 154 | 81 | 14 | 154 | 95 |
| asthma no | 504 | 251 | 36 | 504 | 287 |
| asthma yes | 35 | 14 | 3 | 35 | 17 |
| taste no | 380 | 180 | 25 | 380 | 205 |
| taste yes | 110 | 44 | 11 | 110 | 55 |
| smell no | 381 | 181 | 25 | 381 | 206 |
| smell yes | 109 | 42 | 11 | 109 | 53 |
| mod | 244 | 122 | 21 | 244 | 143 |
| ser | 90 | 46 | 1 | 90 | 47 |
| mild | 124 | 59 | 9 | 124 | 68 |
| crit | 82 | 38 | 8 | 82 | 46 |
| mild+mod | 368 | 181 | 30 | 368 | 211 |
| ser+crit | 172 | 84 | 9 | 172 | 93 |
| death no | 450 | 227 | 31 | 450 | 258 |
| death yes | 90 | 38 | 8 | 90 | 46 |

| | MAF | HWE | $Khi^2$ |
|---|---|---|---|
| all | 0.2032 | 0.8280 | |
| male | 0.2095 | 0.9992 | 0.3461 |
| female | 0.1959 | 0.6926 | |
| diab no | 0.1955 | 0.9265 | 0.3079 |
| diab yes | 0.2204 | 0.8976 | |
| T1DM | 0.2647 | 0.9854 | 0.3013 |
| T2DM | 0.2189 | 0.8682 | 0.3723 |
| asthma no | 0.2042 | 0.8985 | 0.6014 |
| asthma yes | 0.1923 | 0.8162 | |
| taste no | 0.1966 | 0.9091 | 0.6837 |
| taste yes | 0.2000 | 0.3698 | |
| smell no | 0.1968 | 0.9178 | 0.5733 |
| smell yes | 0.1975 | 0.3159 | |
| mod | 0.2119 | 0.7487 | |
| ser | 0.1752 | 0.2852 | |
| mild | 0.2005 | 0.9236 | |
| crit | 0.2109 | 0.7103 | |
| mild+mod | 0.2081 | 0.6930 | 0.7050 |
| ser+crit | 0.1925 | 0.9737 | |
| death no | 0.2041 | 0.9704 | 0.5604 |
| death yes | 0.1985 | 0.6364 | |

### RAVER1

| n | wt | hetero | homo | wt | MV |
|---|---|---|---|---|---|
| all | 762 | 73 | 5 | 762 | 78 |
| male | 407 | 36 | 2 | 407 | 38 |
| female | 355 | 37 | 3 | 355 | 40 |
| diab no | 525 | 44 | 3 | 525 | 47 |
| diab yes | 236 | 29 | 2 | 236 | 31 |
| T1DM | 16 | 2 | 0 | 16 | 2 |
| T2DM | 216 | 27 | 2 | 216 | 29 |
| asthma no | 713 | 69 | 5 | 713 | 74 |
| asthma yes | 48 | 4 | 0 | 48 | 4 |
| taste no | 533 | 44 | 4 | 533 | 48 |
| taste yes | 145 | 17 | 1 | 145 | 18 |
| smell no | 536 | 45 | 4 | 536 | 49 |
| smell yes | 141 | 16 | 1 | 141 | 17 |
| mod | 340 | 37 | 3 | 340 | 40 |
| ser | 128 | 13 | 0 | 128 | 13 |
| mild | 183 | 9 | 0 | 183 | 9 |
| crit | 111 | 14 | 2 | 111 | 16 |
| mild+mod | 523 | 46 | 3 | 523 | 49 |
| ser+crit | 239 | 27 | 2 | 239 | 29 |
| death no | 645 | 58 | 3 | 645 | 61 |
| death yes | 117 | 15 | 2 | 117 | 17 |

| | MAF | HWE | Khi$^2$ |
|---|---|---|---|
| all | 0.0494 | 0.4798 | |
| male | 0.0449 | 0.7846 | 0.4289 |
| female | 0.0544 | 0.6126 | |
| diab no | 0.0437 | 0.5913 | 0.1148 |
| diab yes | 0.0618 | 0.8251 | |
| T1DM | 0.0556 | 0.9710 | 0.6613 |
| T2DM | 0.0633 | 0.8085 | 0.1026 |
| asthma no | 0.0502 | 0.4577 | 0.6808 |
| asthma yes | 0.0385 | 0.9608 | |
| taste no | 0.0448 | 0.3849 | 0.2697 |
| taste yes | 0.0583 | 0.9269 | |
| smell no | 0.0453 | 0.3980 | 0.3501 |
| smell yes | 0.0570 | 0.9109 | |
| mod | 0.0566 | 0.6303 | |
| ser | 0.0461 | 0.8546 | |
| mild | 0.0234 | 0.9474 | |
| crit | 0.0709 | 0.6208 | |
| mild+mod | 0.0455 | 0.6275 | 0.2940 |
| ser+crit | 0.0578 | 0.7750 | |
| death no | 0.0453 | 0.7327 | 0.1390 |
| death yes | 0.0709 | 0.6436 | |

### MUC5B

| n | wt | hetero | homo | wt | MV |
|---|---|---|---|---|---|
| all | 710 | 140 | 11 | 710 | 151 |
| male | 379 | 71 | 8 | 379 | 79 |
| female | 331 | 69 | 3 | 331 | 72 |
| diab no | 480 | 99 | 9 | 480 | 108 |
| diab yes | 229 | 41 | 2 | 229 | 43 |
| T1DM | 13 | 5 | 0 | 13 | 5 |
| T2DM | 212 | 36 | 2 | 212 | 38 |
| asthma no | 663 | 134 | 10 | 663 | 144 |
| asthma yes | 46 | 6 | 1 | 46 | 7 |
| taste no | 498 | 93 | 6 | 498 | 99 |
| taste yes | 137 | 27 | 3 | 137 | 30 |
| smell no | 500 | 93 | 8 | 500 | 101 |
| smell yes | 134 | 27 | 1 | 134 | 28 |
| mod | 324 | 66 | 3 | 324 | 69 |
| ser | 123 | 16 | 2 | 123 | 18 |
| mild | 153 | 39 | 2 | 153 | 41 |
| crit | 110 | 19 | 4 | 110 | 23 |
| mild+mod | 477 | 105 | 5 | 477 | 110 |
| ser+crit | 233 | 35 | 6 | 233 | 41 |
| death no | 590 | 124 | 8 | 590 | 132 |
| death yes | 120 | 16 | 3 | 120 | 19 |

| | MAF | HWE | $Khi^2$ |
|---|---|---|---|
| all | 0.0941 | 0.6768 | |
| male | 0.0950 | 0.4375 | 0.8122 |
| female | 0.0931 | 0.9781 | |
| diab no | 0.0995 | 0.6406 | 0.3591 |
| diab yes | 0.0827 | 0.9970 | |
| T1DM | 0.1389 | 0.8172 | 0.3126 |
| T2DM | 0.0800 | 0.9735 | 0.2687 |
| asthma no | 0.0954 | 0.7733 | 0.3901 |
| asthma yes | 0.0755 | 0.7690 | |
| taste no | 0.0879 | 0.8946 | 0.6736 |
| taste yes | 0.0988 | 0.7617 | |
| smell no | 0.0907 | 0.6292 | 0.8853 |
| smell yes | 0.0895 | 0.9775 | |
| mod | 0.0916 | 0.9916 | |
| ser | 0.0709 | 0.6655 | |
| mild | 0.1108 | 0.9795 | |
| crit | 0.1015 | 0.3755 | |
| mild+mod | 0.0980 | 0.9774 | 0.1748 |
| ser+crit | 0.0858 | 0.2430 | |
| death no | 0.0970 | 0.9397 | 0.1903 |
| death yes | 0.0791 | 0.4294 | |

### UGT2A1

| n | wt | hetero | homo | wt | MV |
|---|---|---|---|---|---|
| all | 319 | 392 | 148 | 319 | 540 |
| male | 159 | 218 | 83 | 159 | 301 |
| female | 160 | 174 | 65 | 160 | 239 |
| diab no | 221 | 262 | 102 | 221 | 364 |
| diab yes | 98 | 130 | 45 | 98 | 175 |
| T1DM | 8 | 7 | 3 | 8 | 10 |
| T2DM | 89 | 121 | 42 | 89 | 163 |
| asthma no | 299 | 366 | 142 | 299 | 508 |
| asthma yes | 20 | 26 | 5 | 20 | 31 |
| taste no | 238 | 253 | 104 | 238 | 357 |
| taste yes | 50 | 85 | 31 | 50 | 116 |
| smell no | 237 | 253 | 108 | 237 | 361 |
| smell yes | 51 | 84 | 27 | 51 | 111 |
| mod | 148 | 173 | 70 | 148 | 243 |
| ser | 53 | 59 | 28 | 53 | 87 |
| mild | 63 | 99 | 32 | 63 | 131 |
| crit | 55 | 61 | 18 | 55 | 79 |
| mild+mod | 211 | 272 | 102 | 211 | 374 |
| ser+crit | 108 | 120 | 46 | 108 | 166 |
| death no | 267 | 322 | 131 | 267 | 453 |
| death yes | 52 | 70 | 17 | 52 | 87 |

| | MAF | HWE | $Khi^2$ |
|---|---|---|---|
| all | 0.4005 | 0.5900 | |
| male | 0.4174 | 0.9276 | 0.0940 |
| female | 0.3810 | 0.5694 | |
| diab no | 0.3983 | 0.5342 | 0.5955 |
| diab yes | 0.4029 | 0.9928 | |
| T1DM | 0.3611 | 0.8964 | 0.5660 |
| T2DM | 0.4067 | 0.9984 | 0.4989 |
| asthma no | 0.4027 | 0.5173 | 0.7564 |
| asthma yes | 0.3529 | 0.8378 | |
| taste no | 0.3874 | 0.2016 | 0.0203 |
| taste yes | 0.4428 | 0.9427 | |
| smell no | 0.3921 | 0.1524 | 0.0578 |
| smell yes | 0.4259 | 0.8625 | |
| mod | 0.4003 | 0.5497 | |
| ser | 0.4107 | 0.5577 | |
| mild | 0.4201 | 0.8966 | |
| crit | 0.3619 | 0.9931 | |
| mild+mod | 0.4068 | 0.8221 | 0.3439 |
| ser+crit | 0.3869 | 0.6692 | |
| death no | 0.4056 | 0.3900 | 0.9418 |
| death yes | 0.3741 | 0.8188 | |

**Supplementary Figure 1. Allele frequencies of the minor variants (MAF) in the hospitalized COVID-19 patients with a severe disease, compared to the MAF values in the general population.**

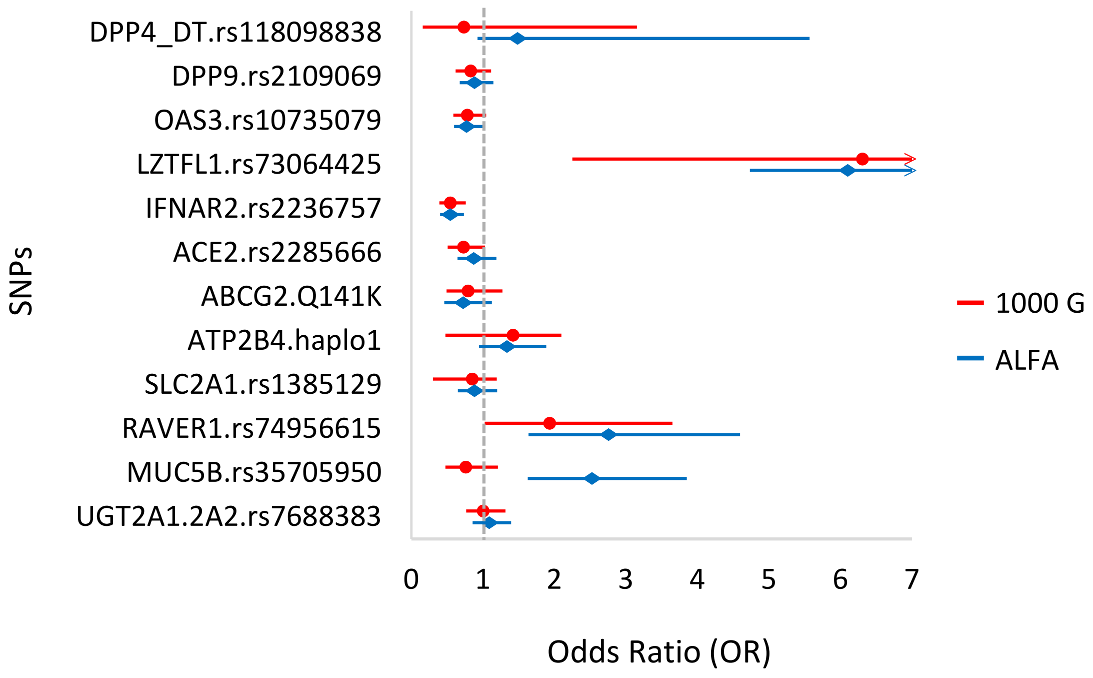


| | Ref.: 1000Genomes (EUR) | | Ref.: ALFA (EUR) | |
|---|---|---|---|---|
| | p-value | OR (95% CI) | p-value | OR (95% CI) |
| **DPP4_DT.rs118098838** | >0.9999 | 0.74 (0.16-3.15) | 0.3964 | 1.49 (0.92-5.57) |
| **DPP9.rs2109069** | 0.2412 | 0.83 (0.62-1.12) | 0.3621 | 0.88 (0.68-1.15) |
| **OAS3.rs10735079** | 0.0999 | 0.78 (0.59-1.05) | 0.0509 | 0.77 (0.60-1.00) |
| **LZTFL1.rs73064425** | <0.0001 | 6.31 (2.25-8.77) | <0.0001 | 6.10 (4.73-7.86) |
| **IFNAR2.rs2236757** | 0.0003 | 0.55 (0.39-0.76) | <0.0001 | 0.55 (0.40-0.74) |
| **ACE2.rs2285666** | 0.0893 | 0.73 (0.51-1.03) | 0.4517 | 0.88 (0.64-1.19) |
| **ABCG2.Q141K** | 0.4072 | 0.79 (0.49-1.27) | 0.1642 | 0.72 (0.46-1.13) |
| **ATP2B4.haplo1** | 0.1011 | 1.42 (0.48-2.10) | 0.0968 | 1.34 (0.95-1.89) |
| **SLC2A1.rs1385129** | 0.3993 | 0.85 (0.30-1.20) | 0.4983 | 0.89 (0.65-1.20) |
| **RAVER1.rs74956615** | 0.0596 | 1.94 (1.03-3.65) | 0.0008 | 2.76 (1.64-4.60) |
| **MUC5B.rs35705950** | 0.3111 | 0.77 (0.48-1.21) | 0.0002 | 2.53 (1.63-3.86) |
| **UGT2A1.2A2.rs7688383** | >0.9999 | 1.01 (0.77-1.32) | 0.4868 | 1.09 (0.86-1.40) |

**Supplementary Figure 2. Allele frequencies of the minor variants (MAF) in the hospitalized COVID-19 patients with an anamnesis of type 2 diabetes, compared to the MAF values in the general population**. *This analysis included 252 diabetic patients.*

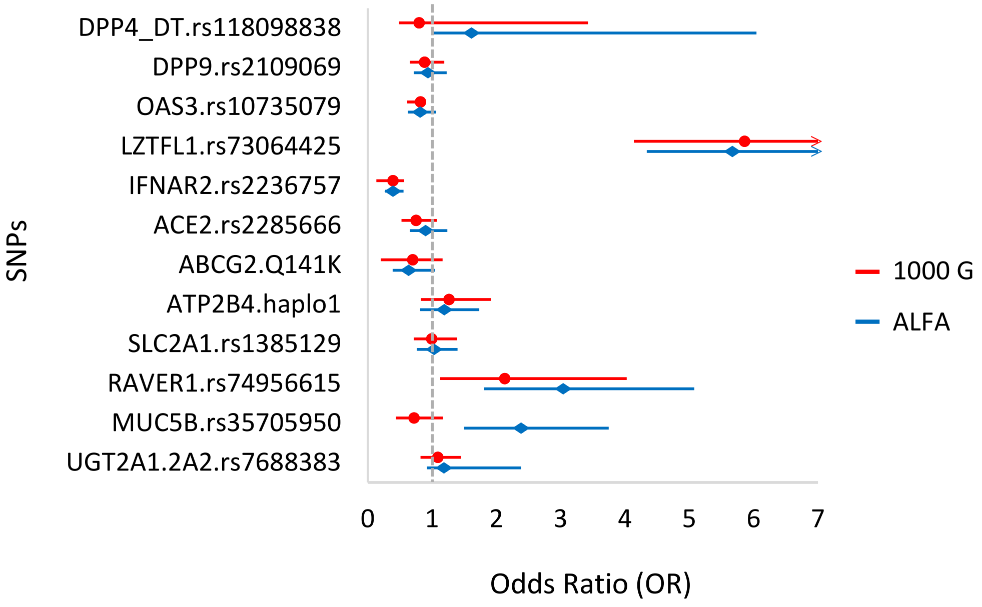


| | Ref.: 1000Genomes (EUR) | | Ref.: ALFA (EUR) | |
|---|---|---|---|---|
| | p-value | OR (95% CI) | p-value | OR (95% CI) |
| **DPP4_DT.rs118098838** | >0.9999 | 0.8 (0.49-3.42) | 0.3594 | 1.61 (1.00-6.05) |
| **DPP9.rs2109069** | 0.4484 | 0.88 (0.66-1.19) | 0.6818 | 0.94 (0.71-1.23) |
| **OAS3.rs10735079** | 0.2097 | 0.82 (0.61-0.61) | 0.1314 | 0.81 (0.62-1.06) |
| **LZTFL1.rs73064425** | <0.0001 | 5.86 (4.14-8.31) | <0.0001 | 5.67 (4.34-7.38) |
| **IFNAR2.rs2236757** | <0.0001 | 0.39 (0.13-0.57) | <0.0001 | 0.39 (0.27-0.56) |
| **ACE2.rs2285666** | 0.1374 | 0.75 (0.52-1.08) | 0.5828 | 0.90 (0.66-1.24) |
| **ABCG2.Q141K** | 0.2156 | 0.70 (0.20-1.17) | 0.0759 | 0.64 (0.39-1.04) |
| **ATP2B4.haplo1** | 0.3003 | 1.26 (0.83-1.92) | 0.3554 | 1.19 (0.82-1.73) |
| **SLC2A1.rs1385129** | >0.9999 | 0.99 (0.71-1.39) | 0.8158 | 1.03 (0.76-1.40) |
| **RAVER1.rs74956615** | 0.0326 | 2.13 (1.13-4.03) | 0.0003 | 3.04 (1.81-5.08) |
| **MUC5B.rs35705950** | 0.2424 | 0.72 (0.44-1.17) | 0.0008 | 2.39 (1.50-3.75) |
| **UGT2A1.2A2.rs7688383** | 0.5632 | 1.09 (0.82-1.45) | 0.2093 | 1.18 (0.92-2.39) |

**Supplementary Figure 3. Allele frequencies of the minor variants (MAF) in the hospitalized COVID-19 patients with or without an anamnesis of loss of taste or smell, compared to the MAF values in the general population.**

**Panel A:** *This analysis included 168 patients with loss of taste*

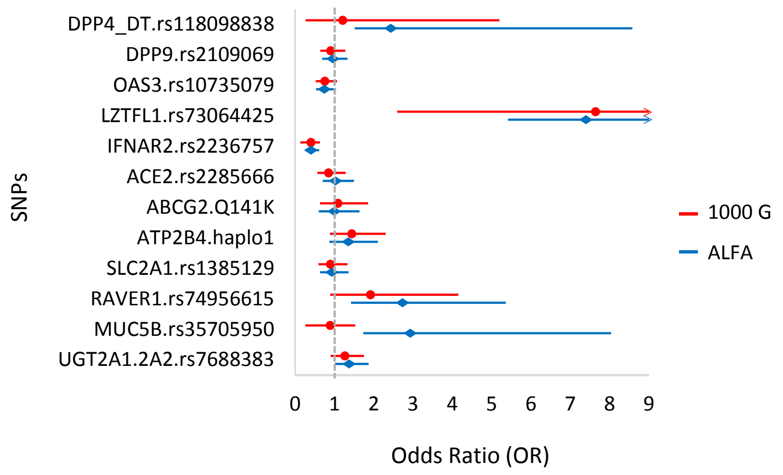


**Panel B. Loss of smell** - *this analysis included 164 patients with loss of smell*

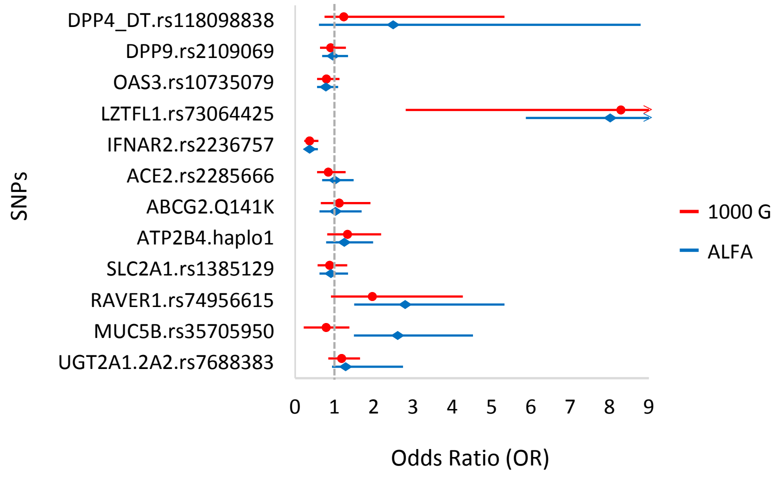


**Supplementary Figure 4. Additional information for the potential correlations of the anamnestic, clinical and genetic data obtained.** *Allele frequencies of the minor variants (MAF) in the hospitalized COVID-19 patients with an anamnesis of asthma, compared to the MAF values in the general population.*

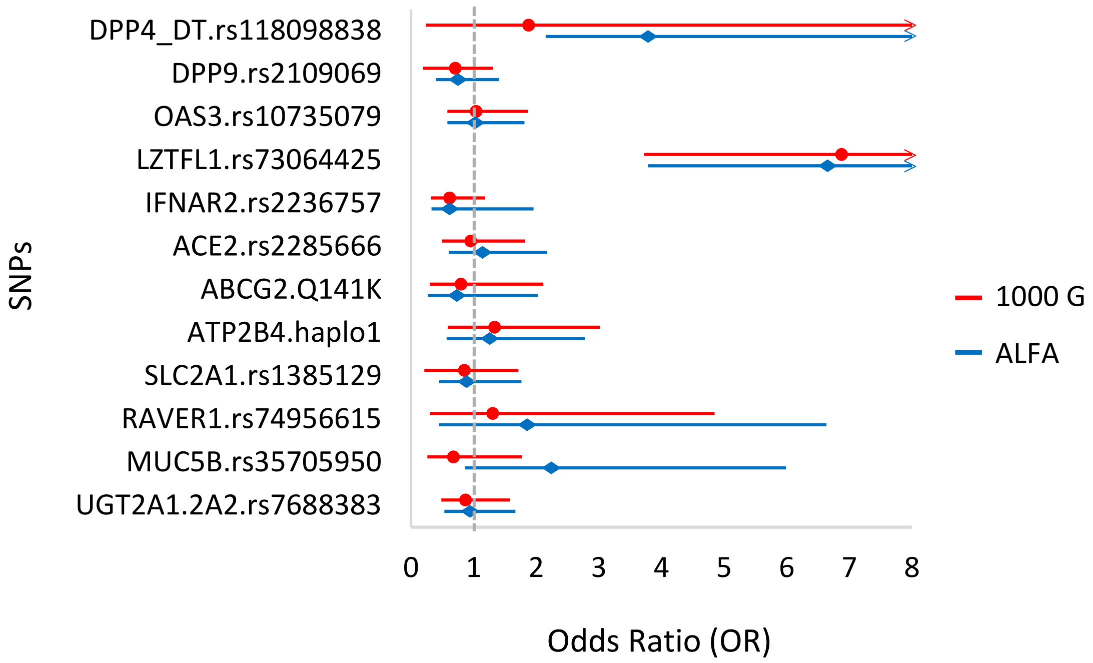


| Gene polymorphism | Ref.: 1000Genomes (EUR) | | Ref.: ALFA (EUR) | |
|---|---|---|---|---|
| | p-value | OR (95% CI) | p-value | OR (95% CI) |
| **DPP4_DT.rs118098838** | 0.4390 | 1.88 (0.24-11.23) | 0.2381 | 3.78 (2.15-21.17) |
| **DPP9.rs2109069** | 0.3594 | 0.70 (0.19-1.30) | 0.4530 | 0.75 (0.40-1.40) |
| **OAS3.rs10735079** | 0.8826 | 1.04 (0.58-1.87) | >0.9999 | 1.03 (0.58-1.81) |
| **LZTFL1.rs73064425** | <0.0001 | 6.87 (3.73-12.62) | <0.0001 | 6.65 (3.79-11.72) |
| **IFNAR2.rs2236757** | 0.1683 | 0.61 (0.32-1.18) | 0.1782 | 0.61 (0.32-1.95) |
| **ACE2.rs2285666** | >0.9999 | 0.95 (0.49-1.82) | 0.7325 | 1.14 (0.60-2.18) |
| **ABCG2.Q141K** | >0.9999 | 0.80 (0.30-2.11) | 0.8176 | 0.73 (0.26-2.02) |
| **ATP2B4.haplo1** | 0.4864 | 1.33 (0.59-3.02) | 0.5093 | 1.25 (0.57-2.78) |
| **SLC2A1.rs1385129** | 0.7329 | 0.85 (0.21-1.72) | 0.8656 | 0.88 (0.44-1.76) |
| **RAVER1.rs74956615** | 0.6681 | 1.30 (0.30-4.85) | 0.3018 | 1.86 (0.45-6.63) |
| **MUC5B.rs35705950** | 0.6462 | 0.68 (0.26-1.78) | 0.1168 | 2.24 (0.86-5.99) |
| **UGT2A1.2A2.rs7688383** | 0.7682 | 0.87 (0.48-1.58) | 0.8854 | 0.94 (0.53-1.67) |
